## Supplementary material for "Access to HIV-prevention in female sex workers in Ukraine between 2009 and 2017: coverage, barriers and facilitators": S1 Appendix. Literature review method, protocol and results

### Supporting Information S1: Literature Review Method, Protocol and Results

### Literature Review Method

The aim of the literature review was to identify, review and summarise the global literature exploring the access that FSW have to HIV prevention, in order to develop a conceptual framework (objective 1). The keywords, “prevent*” AND (“sex work” OR SW OR “sex worker”) AND female AND (“human immunodeficiency virus” OR HIV OR HIV/AIDS) were used to search PubMed. We considered both quantitative and qualitative studies as well as literature reviews that were published in the English language. There was no limitation on publication date. We extracted data on outcome measure, magnitude and direction of effect, confidence intervals and statistical significance. Additionally, we employed snowball searching from identified references to further identify studies informative for the development of the conceptual framework. Both the protocol and the results are available the protocol in S1 Protocol and table in S2 Table, respectively.

### Literature Review Protocol

**Eligibility criteria:**

**Years considered**: no limit (any until 2019)

**Language**: limit to English language

**Publication status**: Empirical studies

I**nformation sources:**

Databases used**:** PubMed

Date of searches**:** 05.02.2019

Contact to study authors to identify additional studies**:** None

Date last searched: 10.02.2019

**Search terms:**

Prevent*

AND

(“Sex work” OR SW OR “Sex worker”) AND female

AND

“Human Immunodeficiency Virus” OR HIV OR HIV/AIDS

- All key terms limited to Title/Abstract

**Study selection:**

1. Titles and abstracts were screened and full text articles were obtained from all selected abstracts.
2. Full-text articles were assessed for eligibility to be included in final study selection, including
   a. Quantitative
   b. Qualitative
   c. Systematic literature reviews

**Data collection process:**

The following data was extracted from full-text articles;

Study identification:

- Author(s)
- Year of publication
- Type of publication

Outcome:

- Outcome measure
- Magnitude of effect
- Direction of effect
- Confidence levels
- Statistical significance
- Only outcomes of statistical significance that were included in, when applicable, a multivariate regression analysis

Other information:

- Definition of prevention service
- Definition of sex work

**Snowball searching**

1. All full-text articles included in final study selection were scanned for relevant references
2. These references were then hand searched and included in final study selection

### Literature Review Results

| **Author(s)** | **Year** | **Methods** | **Country** | **Determinants of HIV prevention access** | **Direction of effect** | **Statistical significance** | **95% CI** | **Definition of prevention** | **Definition of sex work** |
| --- | --- | --- | --- | --- | --- | --- | --- | --- | --- |
| Asadi-AliAbadi | 2018 | Qualitative | Iran | Fear of being infected with HIV | Decrease in access to HIV prevention | N/A | N/A | Not defined | Not defined |
|  |  |  |  | Lack of confidentiality from staff at the HCF |  |  |  |  |  |
|  |  |  |  | Inappropriate location of HCF [not integrated] |  |  |  |  |  |
|  |  |  |  | Drug addiction |  |  |  |  |  |
|  |  |  |  | Lack of knowledge due to advertising |  |  |  |  |  |
| Biradavolu Et al | 2012 | Qualitative | India | NGO provide free condoms | Increase chance of accessing outreach program | N/A | N/A | Not defined | Not defined |
|  |  |  |  | Stigma associated with HIV | Reduced chance of accessing outreach program |  |  |  |  |
|  |  |  |  | Fear of being associated as a SW |  |  |  |  |  |
| Chanda et al | 2017 | Qualitative | Zambia | Fear of self-stigma | Decrease likelihood of accessing HIV testing | N/A | N/A | Not defined | Not defined |
|  |  |  |  | Lack of confidentiality at HCF |  |  |  |  |  |
|  |  |  |  | Lack of information about HIV/AIDS |  |  |  |  |  |
|  |  |  |  | Lack of confidentiality at intrapersonal (amongst friends and wider community) level |  |  |  |  |  |
|  |  |  |  | Intimate partner violence |  |  |  |  |  |
|  |  |  |  | HIV stigma within HCF |  |  |  |  |  |
|  |  |  |  | Sex worker stigma |  |  |  |  |  |
|  |  |  |  | Mandatory HIV testing at HCF |  |  |  |  |  |
|  |  |  |  | Integrated HIV prevention at HCF |  |  |  |  |  |
|  |  |  |  | Inappropriate opening hours |  |  |  |  |  |
|  |  |  |  | Knowledge of risks posed in SW | Increased likelihood of accessing HIV testing |  |  |  |  |
|  |  |  |  | Pregnancy |  |  |  |  |  |
|  |  |  |  | Experiences with family living with or dying from HIV |  |  |  |  |  |
|  |  |  |  | Non-judgemental experiences with health care providers |  |  |  |  |  |
|  |  |  |  | Encouragement from non-paying sexual partners to be HIV tested |  |  |  |  |  |
|  |  |  |  | Encouragement from fellow FSW to be HIV tested |  |  |  |  |  |
|  |  |  |  | Encouragement from friends [not FSW] to be HIV tested |  |  |  |  |  |
|  |  |  |  | Exposure to anti-stigma practices (e.g. skits) |  |  |  |  |  |
|  |  |  |  | Exposure to peer education |  |  |  |  |  |
| Decker et al | 2016 | Quantitative | Cameroon | Gender-based violence | Increased fear of accessing HIV health services (2,25 aOR) | p < 0.0001 | [1,16-3,16] | Not defined | “Selling sex within the past 12 months resulted in more than half their income” |
|  |  |  |  |  | Increased likelihood of being mistreated in a health centre (aOR 1.66) | p < 0.003 | [1.01-2.73] |  |  |
| Decker et al | 2013 | Mixed methods | Russia | Participation in HIV intervention | Increase chance of HIV testing | <0.0001 | N/A | “Having either talked with an outreach worker or attended a Globus clinic” | "Having traded sex for money, drugs or shelter in the past three months” |
|  |  |  |  |  | Increased uptake of condoms |  |  |  |  |
|  |  |  |  |  | Increase complete knowledge of HIV |  |  |  |  |
|  |  |  |  | Confidentiality of staff | Increased participation in HIV program | N/A | N/A |  |  |
|  |  |  |  | Free prevention services (HIV testing) |  |  |  |  |  |
|  |  |  |  | Respectful staff |  |  |  |  |  |
|  |  |  |  | Presence of HIV information |  |  |  |  |  |
|  |  |  |  | Stigma | Decrease uptake of HIV prevention services offered in government-run programs |  |  |  |  |
|  |  |  |  | Lack of local registration papers |  |  |  |  |  |
|  |  |  |  | Police crack-downs |  |  |  |  |  |
|  |  |  |  | Financial limitation |  |  |  |  |  |
| Duby et al | 2018 | Qualitative | South Africa | Stigma within the HCF | Reduced likelihood to visiting a HIV prevention clinic | N/A | N/A | Not defined | Not defined |
|  |  |  |  | Self-Stigma |  |  |  |  |  |
|  |  |  |  | Lack of access to clean drug injecting equipment |  |  |  |  |  |
| Fobosi et al | 2017 | Qualitative | South Africa | Ability to identify risks involved in SW | Increased chance of accessing roadside wellness clinics | N/A | N/A | Not defined | “Reporting having had sex in exchange for goods or money during the past 3 months” |
|  |  |  |  | Appropriate location of the clinic [close-by place of work] |  |  |  |  |  |
|  |  |  |  | Services free of charge |  |  |  |  |  |
|  |  |  |  | Appropriate opening hours of HCF |  |  |  |  |  |
|  |  |  |  | Positive experience with non-judgemental HCF staff |  |  |  |  |  |
| Grayman et al | 2005 | Mixed Methods | Vietnam | History of rehabilitation | Increase likelihood of HIV testing (aOR = 18.40) | <0.001 | [7,40-45.70] | Not defined | Not defined |
| Harawa & Bingham | 2009 | Mixed methods | USA | Hispanic ethnicity | Increase exposure to passive prevention (aOR= 10.14) | N/A | Mistake in article | Passive prevention one of: having observed prevention messages or information in various settings and types of media (including television, Internet, billboards, radio, magazines/newspapers, health centres, clubs/bars, and buses/bus stops), having received printed HIV materials, and having received free condoms Active prevention = contacting an HIV hotline, discussing prevention with an outreach worker, participating in a prevention group session, role-playing safer sex negotiation, exchanging needles for drug injection, receiving HIV testing, receiving an HIV risk reduction plan with HIV testing, and currently participating in drug or alcohol abuse treatment | “Had participated in exchanged sex in the past 12 months” |
|  |  |  |  | Married or cohabitating | Decrease exposure to passive prevention (aOR= 0.18) |  | [0.03-0.98] |  |  |
|  |  |  |  | Any binge drinking [= 5 or more drinks in a single day] | Increase exposure to passive prevention (aOR= 15.18) |  | [1.31-175.82] |  |  |
|  |  |  |  | Any illicit drug use | Increase exposure to passive prevention (aOR= 6.26) |  | [1.19-35.48] |  |  |
|  |  |  |  | 18-24 years | Decrease exposure to active prevention (aOR= 0.26) |  | [0.09-0.87] |  |  |
| Huang et al | 2015 | Quantitative | China | Affluent workplace | Increase likelihood of HIV testing (OR= 4.66) | p < 0.10 | [1.86-11.50] | Prevention = HIV testing | “Reported having received money or other financial benefits in exchange for sexual services during the past 6 months” |
|  |  |  |  | Junior high school or less education level | Increase likelihood of HIV testing (OR = 2.22) |  | [1.01-4.88] |  |  |
|  |  |  |  | Knowledge of where to get HIV test | Increase likelihood of HIV testing (OR = 3.00) |  | [1.42-6.34] |  |  |
|  |  |  |  | STI and HIV/AIDS-related knowledge [above average in population] | Increase likelihood of HIV testing (aOR= 1.36) |  | [1.02- 1.81] |  |  |
|  |  |  |  | Consistent condom use with boyfriend or lover in the past 6 months | Increase likelihood of HIV testing (aOR= 2.75) |  | [1.29 – 5.84] |  |  |
| Izulla et al | 2012 | Mixed methods | Kenya | Presence of a non-paying partner | Decreased likelihood of accessing PEP (OR = 0.52) | p < 0.0001 | [0.39-0.68] | Prevention = PEP | Not defined |
|  |  |  |  | Reporting 100% condom use with casual clients | Increased likelihood of accessing PEP (OR= 1.80) |  | [1.38 – 2-35] |  |  |
|  |  |  |  | Previous HIV test completion | Increased likelihood of accessing PEP (OR = 2.22) |  | [1.45-3.40] |  |  |
| Jayanna et al | 2009 | Qualitative | India | Home based | Increase in STI service uptake | N/A | N/A | Not defined | Not defined |
|  |  |  |  | Street-based | Decrease in STI service uptake |  |  |  |  |
|  |  |  |  | Lodge-based |  |  |  |  |  |
|  |  |  |  | Lack of time to tend to health |  |  |  |  |  |
|  |  |  |  | Inappropriate location of HCF [too far away] |  |  |  |  |  |
|  |  |  |  | High expense of transport to HCF |  |  |  |  |  |
|  |  |  |  | Current unresolved clinical symptoms |  |  |  |  |  |
| Johnston et al | 2017 | Mixed methods [IBBS] | Dominican Republic | Participation in a HIV program (within last 6 months) | Increase chance of testing for HIV (aOR = 2.4) | p < 0.001 | [1.8-3.1] | Access to HIV test | “Engaged in vaginal or anal penetrative sex in exchange for money within the past 6 months” |
|  |  |  |  | Ever had regular health care check ups | Increase chance of testing for HIV (aOR = 2.2) |  | [1.6- 2.9] |  |  |
| Lim et al | 2018 | Qualitative | Singapore | Fear of identity exposure | Decrease | N/A | N/A | Access to health services | Not defined |
|  |  |  |  | Fear of police arrest |  |  |  |  |  |
|  |  |  |  | Stigmatisation |  |  |  |  |  |
|  |  |  |  | Cost of prevention |  |  |  |  |  |
|  |  |  |  | Language difference |  |  |  |  |  |
| Maher et al | 2015 | Qualitative | Cambodia | Prohibition of sex work | Decrease access to health services | N/A | N/A | Not defined | “reported transactional sex [sex in exchange for money, goods, services or drugs] in the Previous three months” |
| Mergenova et al | 2018 | Mixed methods | Kazakhstan | High stigma from health care staff | Decrease in HIV prevention participation | N/A | N/A | Microfinancing [vocational training and financial literacy classes] | “Reporting providing sex in return for money, goods, drugs, or services in the prior 90 days” |
|  |  |  |  | Substance abuse |  |  |  |  |  |
|  |  |  |  | Cost of transport to prevention |  |  |  |  |  |
|  |  |  |  | Incompatible working hour/ inappropriate opening hours |  |  |  |  |  |
| Morales-Miranda et al | 2014 | Quantitative [IBBS] | Guatemala | History of practicing SW in a foreign country | Decrease attendance to health clinic (aOR = 0.75) |  | [0.57-0.98] | Appearing more than once to a prevention site within a 12 month period of initial visit [having received HIV counselling, promotion of condom use, physical evaluation, HIV and syphilis testing] | “Reported exchanging sex for money in the last 12 months” |
|  |  |  |  | Current HIV diagnosis | Decrease attendance to health clinic (aOR= 0.39) |  | [0.18-0.85] |  |  |
| Muñoz, Adedimeji & Alawode | 2010 | Qualitative | Nigeria | High expense of prevention | Decrease likelihood of testing for HIV | N/A | N/A | HIV testing (time period not specified) | “Self identified as a sex worker” |
|  |  |  |  | Stigma within FSW community |  |  |  |  |  |
|  |  |  |  | Lack of confidentiality of health care staff |  |  |  |  |  |
| Ndori-Mharadze et al | 2018 | Mixed methods | Zimbabwe | Free prevention | Increase engagement in HIV prevention clinics | N/A | N/A | Testing in the last 6 months amongst HIV negative FSW” and “self reported service uptake including contact with peer educators, visits to the clinic, testing behaviour and perceived social cohesion” | “Reported exchanging sex for money in the past 30 days” |
|  |  |  |  | Staff considered welcoming and accepting |  |  |  |  |  |
|  |  |  |  | Availability of comprehensive vaginal examinations available |  |  |  |  |  |
|  |  |  |  | Reliable supply of medication |  |  |  |  |  |
|  |  |  |  | Not prioritising ones health | Decrease engagement in HIV prevention clinics |  |  |  |  |
|  |  |  |  | Being lazy |  |  |  |  |  |
|  |  |  |  | Wanting to hide FSW status |  |  |  |  |  |
| Nnko et al | 2019 | Systematic literature review | Sub-Saharan Africa | Discrimination in FSW community | Decrease access likelihood of HTC | N/A | N/A | HIV testing and counselling (HTC) | “females who exchange sex for money or other commodities either regularly or occasionally” |
|  |  |  |  | Inappropriate policies (e.g. partner presence, demand for identity cards) |  |  |  |  |  |
|  |  |  |  | Opportunistic costs (e.g. bribery) |  |  |  |  |  |
|  |  |  |  | Expense of services |  |  |  |  |  |
|  |  |  |  | Transport costs to HCF too expensive |  |  |  |  |  |
|  |  |  |  | Long waiting times at HCF |  |  |  |  |  |
|  |  |  |  | Lack of privacy whilst testing at a HCF |  |  |  |  |  |
|  |  |  |  | Stigma surrounding HIV |  |  |  |  |  |
|  |  |  |  | HCF too close to place of residence |  |  |  |  |  |
|  |  |  |  | Perception of being personally at risk | Increase likelihood of HTC |  |  |  |  |
|  |  |  |  | HCF too far away to place of residence |  |  |  |  |  |
|  |  |  |  | Encouragement from FSWs peers to test for HIV |  |  |  |  |  |
|  |  |  |  | Trained FSWs assist HTC |  |  |  |  |  |
|  |  |  |  | History of HTC |  |  |  |  |  |
|  |  |  |  | Appropriate opening hours of HCF |  |  |  |  |  |
|  |  |  |  | Awareness of the existence and importance of HTC (e.g. moonlight hours) |  |  |  |  |  |
| Papworth et al | 2015 | Quantitative | Burkina Faso | Motherhood | Decreased likelihood of reporting difficulty when accessing health services (aOR = 0.15) |  | [0.34 -0.67] | “Non-barrier contraception, HIV testing” | Not defined |
|  |  |  |  |  | Increased likelihood of ever testing for HIV (aOR = 1.89) | <0.001 | [1.55-2.31] |  |  |
| Prakash et al | 2016 | Secondary quantitative analysis | India | Gender-based violence | 10.3% less likely to access STI treatment in an NGO | p ≤ 0.05 | N/A | “Participants sought treatment/ medicine / advice for the last STI symptom from the government or private hospitals/ clinics, traditional health, medical shops or through the help of NGO-run-clinics” | “who sold sex in exchange for cash at least once in the last one month” |
| Qiao et al | 2014 | Quantitative | China | Fear of breaching FSW status | Decrease chance of taking a HIV test (aOR = 0.514) | p<0.001 | [0.36-0.73] | “Participation in any of the three HIV prevention activities including condom distribution and/ or voluntary counselling and testing [VCT], community-based methadone maintenance treatment program and/ or needle exchange program and peer HIV/AIDS education |  |
|  |  |  |  | Working in venues with less mobility | Increase chance of having taken a HIV test (aOR = 1.750) | p <0.01 | [1.241-2.467] |  |  |
|  |  |  |  | Younger clientele | Decrease chance of having taken a HIV test (aOR= 0.662) | p<0.05 | [0.441 - 0.993] |  |  |
|  |  |  |  | High awareness of HIV | Increase chance of having taken a HIV test (aOR = 2.815) | p<0.001 | [1.964 - 4.036] |  |  |
|  |  |  |  | Having been arrested by police | Increase chance of being test for HIV in the past (OR = 0.65) | p<0.05 | [0.492-0.871] |  |  |
|  |  |  |  | Higher annual income | Increase chance of participating in HIV program (aOR= 1.874) | p<0.001 | [1.339 - 2.621] |  |  |
|  |  |  |  | Being involved in sex work for longer than one year | Increase chance of participating in HIV program (aOR = 2.09) | p<0.05 | [1.218-3.585] |  |  |
|  |  |  |  | Lower charge per sexual transaction | Reduced change of participating in HIV program |  |  |  |  |
| Restar et al | 2017 | Qualitative | Kenya | Trustworthiness of health care professional | Increase attendance at HIV prevention facility | N/A | N/A | Not defined | “Had vaginal or anal intercourse at least once in the last three months with a paying client that they had met at the venue” |
|  |  |  |  | Overcrowded facilities |  |  |  |  |  |
|  |  |  |  | Confidentiality of HCW |  |  |  |  |  |
|  |  |  |  | Traveling too far for HIV prevention |  |  |  |  |  |
|  |  |  |  | High quality sexual and reproductive health examinations offered at HCF |  |  |  |  |  |
|  |  |  |  | Location too close to home |  |  |  |  |  |
| Rocha-Jiménez et al | 2016 | Qualitative | Guatemala | Fear of being identified as SW | Decreased likelihood of attending HIV services | N/A | N/A | Not defined | Those who exchanged sex for money, drugs or other resources in the last 6 months” |
|  |  |  |  | Lack of confidentiality |  |  |  |  |  |
|  |  |  |  | Attend a private doctor for HIV testing |  |  |  |  |  |
|  |  |  |  | Migrant status |  |  |  |  |  |
|  |  |  |  | Not wanting family to know FSW profession |  |  |  |  |  |
| Scorgie et al | 2012 | Qualitative | Kenya, - Zimbabwe, - Uganda, South Africa | Long waiting times at HCF | Decrease likelihood to HIV prevention access | N/A | N/A | Not defined | “Any agreement between two or more persons in which the objective is exclusively limited to the sexual act and ends with that and which involves preliminary negotiations for a price” |
|  |  |  |  | Inappropriate policies (e.g. only monogamous couples, must bring partner) |  |  |  |  |  |
|  |  |  |  | Self-stigma |  |  |  |  |  |
|  |  |  |  | Unaware of where to access HIV prevention |  |  |  |  |  |
|  |  |  |  | Stigma from HCW |  |  |  |  |  |
|  |  |  |  | Lack of confidentiality from HCW |  |  |  |  |  |
|  |  |  |  | Criminalisation of sex work |  |  |  |  |  |
|  |  |  |  | Having to bribe HCW |  |  |  |  |  |
|  |  |  |  | HCF too far away to place of residence |  |  |  |  |  |
|  |  |  |  | Discrimination from HCW |  |  |  |  |  |
|  |  |  |  | Cost of transport to HCF |  |  |  |  |  |
|  |  |  |  | Shortages of medicine at HCF |  |  |  |  |  |
|  |  |  |  | High user fees |  |  |  |  |  |
|  |  |  |  | Discrimination free-staff |  |  |  |  |  |
| Travasso et al | 2014 | Quantitative | India | Non-cohabiting non-paying partner | Increase chance of being contacted by a peer educator in past one month (aOR= 1.7) | p<0.001 | [1.3-2.1] | "Contact by a peer education in the one month prior to survey” | Defined elsewhere |
|  |  |  |  |  | Increase chance of attending meetings organized by an NGO (aOR= 1.5) | p<0.001 | [1.2-1.8] | Not defined |  |
|  |  |  |  |  | Increased chance of visiting an STI clinic at least twice in last six months (aOR= 1.6) | p <0.001 | [1.3-1.9] | "At least two visits to an STI clinic in the six months prior to survey" |  |
| Tucker et al | 2011 | Qualitative | China | Having a laoxiang (friend from original home town) who encourages HIV testing | Increase likelihood of attending HIV testing | N/A | N/A | STI/HIV testing | “Women who sold sex in the past month for less than five US dollars per client, referred to as low-income sex workers” |
| Wanyenze et al | 2017 | Qualitative | Uganda | Individual level of knowledge and awareness of HIV services | Increase likelihood of access condoms, HTC or STI testing | N/A | N/A | Not defined | Not defined |
|  |  |  |  | Belonging to social network with positive HIV + support, that encouraged testing |  |  |  |  |  |
|  |  |  |  | Fear of breach of confidentiality due to limited privacy at HCF | Decreased likelihood of accessing condoms, HTC or STI testing |  |  |  |  |
|  |  |  |  | Unwelcoming attitude of health workers |  |  |  |  |  |
|  |  |  |  | Discrimination |  |  |  |  |  |
|  |  |  |  | Unfavourable opening hours of HCFs |  |  |  |  |  |
|  |  |  |  | Unfavourable policies (e.g. Requirement of partner presence) of HCF |  |  |  |  |  |
|  |  |  |  | Doubting ones HIV test results based on negative self esteem |  |  |  |  |  |
|  |  |  |  | Fear to know HIV status |  |  |  |  |  |
|  |  |  |  | Fear of stigma |  |  |  |  |  |
|  |  |  |  | Fear that HIV+ status would affect business |  |  |  |  |  |
|  |  |  |  | Belief that health workers may give them drugs that could kill them |  |  |  |  |  |
|  |  |  |  | High mobility of FSW |  |  |  |  |  |
|  |  |  |  | High level of client violence |  |  |  |  |  |
|  |  |  |  | Previous experience of waiting too long in HCF |  |  |  |  |  |
|  |  |  |  | Fear of being identified as FSW |  |  |  |  |  |
|  |  |  |  | Stigma within FSW community |  |  |  |  |  |
|  |  |  |  | Fear of stigma from family |  |  |  |  |  |
|  |  |  |  | Fear of stigma from community |  |  |  |  |  |
|  |  |  |  | Fee of services |  |  |  |  |  |
|  |  |  |  | Expectation of needing to pay HCW extra |  |  |  |  |  |
|  |  |  |  | Low income |  |  |  |  |  |
|  |  |  |  | Transport too expensive |  |  |  |  |  |
|  |  |  |  | Fear of being arrested |  |  |  |  |  |
| Yi et al | 2010 | Quantitative [Cross sectional] | China | Street-based FSW | Decreased likelihood of receiving community health outreach | p< 0.001 | N/A | Exposure to community health outreach | “Transactional sexual exchange for money from clients, regardless of one’s own perception of “sex work”” |
|  |  |  |  |  | Decreased likelihood of testing for HIV in past 12 months | p<0.01 |  | HIV testing in last 12 months |  |
| Zeng et al | 2016 | Qualitative [face-to-face in depth interviews] | China | Lack of knowledge and awareness of HIV prevention services | Decrease likelihood of access to prevention | N/A | N/A | Not defined | "One who conducts sex work on streets, open areas or public venues [e.g. parks or toilets] and exchanged sex for goods or money during the 30 days prior to the study” |
|  |  |  |  | Financial constraints to cover out-of-pocket costs |  |  |  |  |  |
|  |  |  |  | Fear of police arrest |  |  |  |  |  |
|  |  |  |  | Fear of stigma in HCF |  |  |  |  |  |
|  |  |  |  | Discrimination from HCW |  |  |  |  |  |
|  |  |  |  | Distrust in health care workers |  |  |  |  |  |
|  |  |  |  | Smaller HCF | Increase likelihood |  |  |  |  |
|  |  |  |  | Free services |  |  |  |  |  |
|  |  |  |  | Broader sexual health services provided |  |  |  |  |  |
| FSW = Female sex worker; HCF = Health care facility; HCW = Health care worker; HIV = Human immunodeficiency virus; PEP = Postexposure prophylaxis | | | | | | | | | |
