## Supplementary material for "Access to HIV-prevention in female sex workers in Ukraine between 2009 and 2017: coverage, barriers and facilitators": S2 Table. Formation of thematic clusters

### Supporting Information S2: Formation of thematic clusters used to populate the conceptual framework

| **Structural level** | **Sub-level** | **Thematic cluster** | **Specific risk factor** | **Categorisation** |
| --- | --- | --- | --- | --- |
| Macrostructural | Legal | Legal status of FSW | Criminalisation of FSW | Barrier |
|  |  |  | Current incarceration | Barrier |
|  |  |  | History of police arrest | Facilitator |
|  |  |  | Fear of arrest | Barrier |
|  |  |  | Demolition of red-light districts | Barrier |
|  |  |  | Infringement of HCF and NGOs who work with FSW | Barrier |
|  |  | Phrasing of FSW law | Conflation of the term 'sex trafficking' with 'sex work' | Barrier |
|  |  |  | Implementation of brothel rescue-raid police practices to combat sex trafficking | Barrier |
|  |  | Legal support of key-populations | Existence of antidiscrimination laws | Facilitator |
|  |  |  | Violence against FSW formally recorded | Facilitator |
|  |  |  | Active monitoring and oversight of antidiscrimination policy | Facilitator |
|  | Socio-political | Political commitment towards reducing the HIV burden among key populations | Investment in HIV-related programmes for FSW (e.g. legal literacy) | Facilitator |
|  |  | Existence of institutions that exercise and enforce non-discriminatory practices (e.g. hiring of SW) | Presence of institutions that practice antidiscrimination policies (e.g. hiring of SW) | Facilitator |
|  |  | National prevalence of homelessness | Residential instability amongst FSW | Barrier |
|  | Socio-cultural | FSW stigma amongst the general population | Fear of being identified as a FSW | Barrier |
|  |  |  | High level of FSW stigma in the general population | Barrier |
|  |  |  | Internationalised stigma from belief that all FSW are HIV positive | Barrier |
|  |  | HIV stigma amongst the general population | Fear of stigma of being HIV positive | Barrier |
|  |  |  | Fear of receiving a positive HIV test result | Barrier |
|  |  |  | Exposure to anti-stigma interventions e.g. skits normalising FSW | Facilitator |
|  | Geographical | Location of HIV prevention service centres | HCF is too far away from place of residence | Barrier |
|  |  |  | Prevention facility is publicly visible | Barrier |
|  |  |  | HIV prevention centre are *not* integrated in the HCF | Barrier |
|  |  |  | HCF is situated close to place of residence | Facilitator |
|  |  | Regional mobility associated with FSW | High mobility of sex work | Barrier |
|  |  |  | Short term travel to sex hot-spots | Barrier |
|  |  |  | Intra-urban or intra-district mobility | Barrier |
|  |  |  | Does not speak the local language | Barrier |
|  |  |  | Ever having practiced SW in a foreign country | Barrier |
|  |  |  | Not registered locally | Barrier |
|  | Health-related Policy | Accepted behaviour of HCW | Lack of confidentiality exhibited by HCW | Barrier |
|  |  |  | Distrust in HCW e.g. belief that HCW will administer the wrong drugs to deliberately kill a FSW | Barrier |
|  |  |  | High level of stigma exhibited by HCW | Barrier |
|  |  |  | High level of trust in HCW | Facilitator |
|  |  |  | History of discrimination at a HCF e.g. when disclosing sexual practices | Barrier |
|  |  | HCF policies regarding the receipt of HIV prevention | Existence of policies that are insensitive to the FSW community e.g. Abstinence only, monogamous couples only, requirement to bring sexual partner, demand for identity card | Barrier |
|  |  |  | Policies that neglect individual consent e.g. mandatory HIV testing policy | Barrier |
|  |  | HIV prevention also available from NGOs | Poor quality of prevention services provided from NGOs e.g. distribution of condoms that break without the application of lubricant | Barrier |
|  |  |  | Existence of NGOs that provide voluntary prevention services | Facilitator |
|  |  |  | Availability of telephone counselling at NGOs | Facilitator |
|  |  |  | Case management practiced at NGO | Facilitator |
|  |  |  | Existence of NGOs that provide anonymous prevention services | Facilitator |
|  |  | Functional hours of HCF | Appropriate opening hours of HCF e.g. 'moonlight' hours | Facilitator |
|  |  |  | Long waiting time at HCF | Barrier |
|  |  | Available services at HCF | Availability of self-help services e.g. HIV self-tests | Facilitator |
|  |  |  | Perception that only low-quality services are available at HCF e.g. fear of being infected by medical testing utensils at the HCF | Barrier |
|  |  |  | Lack of available services that are tailored to the FSW community e.g. no anal and vaginal examinations | Barrier |
|  |  |  | A lack of harm reduction services offered at HCF e.g. OST | Barrier |
|  |  |  | Insufficient supply of HIV prevention | Barrier |
|  |  |  | Support services for HCW e.g. training about the specific needs of FSWs |  |
|  | Socio - economic | National financial scheme for HIV prevention | Bribes are expected from HCW | Barrier |
|  |  |  | Transportation to HCF is too expensive | Barrier |
|  |  |  | HCF services are free of charge | Facilitator |
|  |  | Level of incentive for women to (re-) enter the workforce | FSW is ones sole source of income | Barrier |
|  |  | National median household income | Higher annual household income (higher than average) | Facilitator |
|  |  |  | Currently attending a private doctor for HIV prevention | Barrier |
| Sex Worker Community Organisation | Community Empowerment | FSW involvement in the development of HIV prevention strategies | Prevention services are carefully tailored to the FSW community | Facilitator |
|  |  |  | History of participation in a HIV program/intervention | Facilitator |
|  |  | FSW engagement with government | Existing dialogue between FSW community and government | Facilitator |
|  |  | FSW engagement with police | Existing dialogue between FSW community and police e.g. training conducted by/with FSW for police about supporting (at least not impeding) FSWs access to health care | Facilitator |
|  | Sex Work Collectivisation | Existence of FSW community organisations (e.g. NGOs or CBOs run by or involving FSWs) | Membership in an NGO that works with FSW community | Facilitator |
|  | Other forms of social and community participation | Collaboration and cooperation between groups of FSW and non-FSW | Participation in non-sex worker community organisation or social networks | Facilitator |
|  | Leadership, education and outreach | Existence of peer-education programs | Exposure to peer- education | Facilitator |
| Work Environment | Physical | Existence of prevention services at work place | Availability of condoms and lubricant at work place | Facilitator |
|  |  | Soliciting setting | Village-based soliciting | Barrier |
|  |  |  | Street-based soliciting | Barrier |
|  |  |  | Working in venues with less client traffic | Facilitator |
|  |  |  | Lodge/hotel-based soliciting | Barrier |
|  |  |  | Home-based soliciting | Facilitator |
|  |  |  | Affluent entertainment venues soliciting | Facilitator |
|  | Social | Local police practices | Pervasive patrolling of HIV facilities | Barrier |
|  |  | Level of gender-based violence at work place | Experience of gender-based violence from police officer in order to avoid arrest | Barrier |
|  |  |  | Experience of gender-based violence from client | Barrier |
|  | Policy | Venue client policies | Client sign in policy | Facilitator |
|  |  |  | Removal of violent clients | Facilitator |
| Interpersonal Dynamic | Sex worker- family* | Family history of HIV* | Being an orphaned child from a parent who has died from HIV* | Facilitator |
|  |  |  | Experience of family living with or dying from HIV* | Facilitator |
|  | Sex worker - Client | Number of clients | High number of clients | Barrier |
|  |  | Condom use | Consistent condom use with casual clients within the past 6 months | Facilitator |
|  | Sex worker -non-paying partner | Gender-based violence | Experience of gender-based violence from an intimate partner | Barrier |
|  |  | Condom use | Consistent condom use with a boyfriend or lover within the past 6 months | Facilitator |
|  |  | Partnership status | Having a non-cohabitation, non-paying partner | Facilitator |
|  |  | Disclosure of occupational status to partner | Non-paying partner is aware of FSW status | Facilitator |
| Sex Worker Individual | Behavioural | Level of education | Junior high school or lower level of education | Facilitator |
|  |  | Work life balance | Lack of time to seek health care | Barrier |
|  |  | History of substance abuse | History of binge drinking | Facilitator |
|  |  |  | Any history of illicit drug use | Facilitator |
|  |  |  | Indifferent outlook to life due to drug addiction, thus not caring about health and wellbeing | Barrier |
|  |  |  | Current drug addiction | Barrier |
|  |  | Duration in sex work | Duration working as a FSW longer than 1 year | Facilitator |
|  |  | Knowledge of HIV | Knowledge of where to get a free HIV test | Facilitator |
|  |  |  | Knowledge of HIV transmission routes | Facilitator |
|  |  |  | Belief that prophylactic use of antibiotics can prevent HIV acquisition | Barrier |
|  |  |  | Belief that FSW can detect clients with symptoms of infection | Barrier |
|  |  |  | Self perception of being high risk for HIV | Facilitator |
|  | Biological | HIV factors | HIV positive status | Barrier |
|  |  |  | History of side effects from HIV medication | Barrier |
|  |  |  | History of being diagnosed with HIV during the first visit to a prevention centre | Barrier |
|  |  | General health status | Unresolved clinical symptoms | Barrier |
|  |  | STI factors | Positive gonorrhoea status | Facilitator |
|  |  | Reproductive status | Motherhood | Facilitator |
|  |  |  | Pregnancy | Facilitator |
|  |  | Age | Age between 18-24 | Barrier |
| Client Individual | Biological | Age | Younger clients | Facilitator |
| HCF = Health Care Facility; HCW = Health Care Worker; OST = Opioid Substitution Therapy | | | | |
| *Specific factors that were identified in the literature review but are not relevant in a Ukrainian context and were therefore not included in the conceptual framework | | | | |
