## Supplementary material for "Access to HIV-prevention in female sex workers in Ukraine between 2009 and 2017: coverage, barriers and facilitators": S3 Table. Categories and specific PP components

### Supporting Information S3: Categories and specific PP components as extracted from the Alliance General Reports from 2008 to 2017

| Category | Prevention Package Components |
| --- | --- |
| **Any form of counselling from a social worker** | - Reproductive and sexual counselling - Social and psychological counselling - Counselling on HIV and STI prevention, to reduce harm of drug and alcohol use, consultations to reduce risky sexual behaviour - Counselling on reproductive and sexual health - Counselling with a focus on evaluating client health status and needs associated with sex work, providing information on safe sexual behaviours and effective ways to prevent the transmission of HIV and STIs etc. - Legal advice - Legal and sociological advice |
| **Any form of specialist medical or legal counselling** | - Professional counselling (psychologist, lawyer, gynaecologist, STI specialist etc.) - Specialist counselling (a social and/or medical work) (not specified further) - Phthisiologist counselling |
| **Some form of peer counselling** | - Peer-driven intervention - Peer driven implementation: repeat coverage - Peer counselling - Peer-driven counselling |
| **Any form of HIV testing and counselling** | - Voluntary HIV *rapid* testing and counselling - Voluntary testing and counselling (VCT) - Assisted testing for HIV |
| **Participation in group-work** | - Group work - Conducting self-help groups which include psychological support and trainings to form safe behaviour skills |
| **Female condoms and training for usage** | - Distribution of female condoms - Training on the use of the female condom |
| **Some form of TB screening** | - Early diagnosis of TB - TB screening survey |
| **Syringe and/or exchange** | - Exchange or delivery of syringes for SW who are IUDs - Syringe distribution and exchange |
| **Training about safe behaviour** | - Involving clients into training activities (not specified further) - Training on safe behaviour development (safe behaviour not defined) |
| **Counteracting violence** | - Counteracting violence (format not specified) - Response to violence against SWs (format not specified) |
| **Distribution of general medication** | - Distribution of medications of general use and providing of first (pre-hospital) aid - Distribution of medications of general use |
| **Any form of professional/ skilled work training** | - Skills training and employment - Vocational training and employment - Professional training and employment - Courses in massage therapy, cosmetology, nutritional science, first pre-medical aid, computer science etc. (*How* not specified) - Sewing and needle work courses |
| **Organisation of leisure activities** | - Organising client’s leisure time - Arrangement of leisure and rest - Social and leisure activities |
| **Basic household services** | - Basic household services (not specified further) - Basic everyday services e.g. shower, laundering, ironing etc. |
| **Tea and meals** | - Tea and meals - Tea and snacks |
| **Childcare and services** | - Temporary child care and help in allocating children to kindergartens - Day-care centres for children - Information and education activities among SW children |
| **Referrals to relevant specialists** | - A system or referrals to relevant specialists - Referral and support in seeking care from service providers to receive diagnostic testing and treatment for HIV, TB and STIs - Referrals to HCF and other targeted projects if necessary (necessary not defined) |
| **Awareness raising and educational material** | - Information regarding the decrease of risky behaviour, HIV and STI prevention, materials indicating medical institution that co-operate with NGOS - Information and educational activities - **Although no explicit item in the prevention package in years 2013 and 2014 were included, a detailed description of the awareness-raising and educational materials distributed to all groups was included in annual reports and therefore were included in this analysis. |
| **Any form of hepatitis C testing** | - Diagnostic and treatment of Hepatitis C - Diagnostics of Hepatitis C |
| **Any form of hepatitis B testing** | - Diagnostics and treatment of Hepatitis B - Diagnostics of Hepatitis B - Assisted testing for HBV |
| **Any form of STI testing** | - Diagnostic and referrals for STI treatment (which STIS not specified) - Diagnostic and treatment of STIS (which treatment exactly, not specified) - Testing for Chlamydia - Testing for gonorrhoea - Assisted testing for syphilis - Examinations for sexually transmitted diseases |
