## Supplementary material for "Access to HIV-prevention in female sex workers in Ukraine between 2009 and 2017: coverage, barriers and facilitators": S4 Table. PP component and corresponding IBBS item

### Supporting Information S4: PP component and corresponding IBBS question for 2009, 2011, 2013, 2015 and 2017

| **2009** | | | | |
| --- | --- | --- | --- | --- |
| **PP component** | **Included in PP** | **Corresponding interview question in IBBS** | **Variable name** | **Definition of variable for analysis** |
| Any form of counselling from a social worker | x | What kind of help or services did you receive from NGOs FOR THE LAST 12 MONTHS?_Legal counselling | p211_10 | Receipt of counselling = p211_10 AND/OR p211_9 |
|  |  | What kind of help or services did you receive from NGOs FOR THE LAST 12 MONTHS?_ HIV / AIDS counselling, sexually transmitted diseases, and ways to prevent them | p211_9 |  |
| Any form of specialist medical or legal counselling | x | What kind of help or services did you receive from NGOs FOR THE LAST 12 MONTHS?_Psychologist consultations | p211_11 | Receipt of specialist counselling = yes to p211_11 |
| Online counselling |  |  |  |  |
| Some form of peer counselling | x | What kind of help or services did you receive from NGOs FOR THE LAST 12 MONTHS? Peer-to-peer counselling (advice given to you by women who provide or previously provided sexual services for you) | p211_12 | Receipt of peer counselling = yes to p211_12 |
| Any form of HIV testing | x | What kind of help or services did you receive from NGOs FOR THE LAST 12 MONTHS?_Free testing for HIV / AIDS | p211_13 |  |
| Participation in group work | x | What kind of help or services did you receive from NGOs FOR THE LAST 12 MONTHS?_Visiting support groups | q211_7 | Participation in group work =yes to p211_7 |
| Male condoms | x | What kind of help or services did you receive from NGOs FOR THE LAST 12 MONTHS?_Receiving condoms | p211_4 | Receipt of male condoms = yes to p211_4 |
| Female condoms and training for usage |  |  |  |  |
| Lubricants | x | - | - | - |
| Some form of TB screening |  |  |  |  |
| Any form of STI testing | x | What kind of help or services did you receive from NGOs FOR THE LAST 12 MONTHS?_Free testing for STDs | p211_14 | Receipt of STI testing = yes to p211_14 AND/OR p211_15 |
|  |  | What kind of help or services did you receive from NGOs FOR THE LAST 12 MONTHS?_Free treatment for sexually transmitted diseases | p211_15 |  |
| Any form of hepatitis C testing |  |  |  |  |
| Any form of hepatitis B testing |  |  |  |  |
| Referral to OST |  |  |  |  |
| Syringe exchange and/or delivery | x | What kind of help or services did you receive from NGOs FOR THE LAST 12 MONTHS?_Syringe exchange | p211_1 | Receipt of syringe exchange yes to = p211_1 |
| Overdose prevention |  |  |  |  |
| Training about safe behaviour |  |  |  |  |
| Case Management |  |  |  |  |
| Counteracting violence |  |  |  |  |
| Distribution of general medication |  |  |  |  |
| Antiseptics | x | What kind of help or services did you receive from NGOs FOR THE LAST 12 MONTHS?_Recovery of disinfection solutions | p211_2 | Receipt of antiseptics = yes to p211_2 |
| Pregnancy tests | x | - | - | - |
| Any form of professional/ skilled work training | x | - | - | - |
| Organisation of leisure activities | x | - | - | - |
| Humanitarian aid | x | - | - | - |
| Basic household services | x | - | - | - |
| Tea and meals | x | - | - | - |
| Childcare and services | x | - | - | - |
| Cosmetologist and hairdresser services | x | - | - | - |
| Intimate hygiene items | x | - | - | - |
| Referrals to relevant specialists | x | - | - | - |
| *Awareness raising and educational material | x | What kind of help or services did you receive from NGOs FOR THE LAST 12 MONTHS?_Receiving information booklets, brochures | p211_5 | Receipt of awareness raising material = yes to p211_5 |
| **2011** | | | | |
| **PP component** | **Included in PP** | **Corresponding interview question in IBBS** | **Variable name** | **Definition of variable for analysis** |
| Any form of counselling from a social worker | x | What kind of help or services did you receive from NGOs FOR THE LAST 12 MONTHS? _ Consultations on HIV / AIDS, sexually transmitted diseases, and ways to prevent them | V199_9 | Receipt of counselling from a social worker = answered yes to V199_9 |
| Any form of specialist medical or legal counselling | x | What kind of help or services did you receive from NGOs FOR THE LAST 12 MONTHS? _ Consultation of a lawyer | V199_10 | Receipt of specialist counselling = answered yes to V199_10 OR V199_11 |
|  |  | What kind of help or services did you receive from NGOs FOR THE LAST 12 MONTHS?_Psychologist consultations | V199_11 |  |
| Online counselling | x | - | - | - |
| Some form of peer counselling | x | What kind of help or services did you receive from NGOs FOR THE LAST 12 MONTHS? _ Peer-to-peer counselling (advice given to women who provide or previously provided sexual services for rewards) | V199_12 | Receipt of peer counselling = answered yes to V199_12 |
| Any form of HIV testing | x | What kind of help or services did you receive from NGOs FOR THE LAST 12 MONTHS?_Free testing for HIV / AIDS | V199_13 | Receipt of HIV= testing answered yes to V199_13 |
| Participation in group work | x | What kind of help or services did you receive from NGOs FOR THE LAST 12 MONTHS? _ Visiting groups of mutual support | V199_7 | Participation in group work = answered yes to V199_7 |
| Male condoms | x | What kind of help or services did you receive from NGOs FOR THE LAST 12 <MONTHS? _ Receiving condoms | V199_4 | Receipt of male condoms = answered yes to V199_4 |
| Female condoms and training for usage | x | - | - | - |
| Lubricants | x | - | - | - |
| Some form of TB screening |  |  |  |  |
| Any form of STI testing | x | How did you treat this disease? TRICHOMONIASIS_I went to NGO | V182_3 | Receipt of STI testing = answered yes to V182_3 OR V167_3 OR V170_3 OR V179_3 OR V164_3 OR V199_14 OR V199_15 |
|  |  | How did you treat this disease? GENITAL HERPES_I went to NGO | V167_3 |  |
|  |  | How did you treat this disease? CHLAMYDIA INFECTION_I went to NGO | V170_3 |  |
|  |  | How did you treat this disease? SYPHILIS_I went to NGO | V179_3 |  |
|  |  | How did you treat this disease? GONORRHEA_I went to NGO | V164_3 |  |
|  |  | What kind of help or services did you receive from NGOs FOR THE LAST 12 MONTHS? _ Free examination for sexually transmitted diseases | V199_14 |  |
|  |  | What kind of help or services did you receive from NGOs FOR THE LAST 12 MONTHS?_Free treatment for sexually transmitted diseases | V199_15 |  |
| Any form of hepatitis C testing | x | - | - | - |
| Any form of hepatitis B testing | x | - | - | - |
| Referral to OST |  |  |  |  |
| Syringe exchange and/or delivery |  |  |  |  |
| Overdose prevention |  |  |  |  |
| Training about safe behaviour |  |  |  |  |
| Case Management |  |  |  |  |
| Counteracting violence | x | - | - | - |
| Distribution of general medication | x | - | - | - |
| Antiseptics | x | What kind of help or services did you receive from NGOs FOR THE LAST 12 MONTHS?_Recovery of disinfection solutions | V199_2 | Receipt of antiseptics = answered yes to V199_2 |
| Pregnancy tests | x | - | - | - |
| Any form of professional/ skilled work training | x | - | - | - |
| Organisation of leisure activities | x | - | - | - |
| Humanitarian aid |  |  |  |  |
| Basic household services | x | - | - | - |
| Tea and meals |  |  |  |  |
| Childcare and services | x | - | - | - |
| Cosmetologist and hairdresser services | x | - | - | - |
| Intimate hygiene items |  | What kind of help or services did you receive from NGOs FOR THE LAST 12 MONTHS? _ Hygiene products | V199_3 | Receipt of intimate hygiene items = answered yes to V199_3 |
| Referrals to relevant specialists | x | - | - | - |
| *Awareness raising and educational material | x | What kind of help or services did you receive from NGOs FOR THE LAST 12 MONTHS?_Receiving information booklets, brochures | V199_5 | Receipt of awareness raising material = answered yes to V199_5 |
| **2013** | | | | |
| **PP component** | **Included in PP** | **Corresponding interview question in IBBS** | **Variable name** | **Definition of variable for analysis** |
| Any form of counselling from a social worker | x | Help from public organizations DURING 12 MONTHS - Advice on less hazardous drug use | q317 | Receipt of social worker counselling = answered yes to q317 OR q318 q316 |
|  |  | Help from public organizations DURING 12 MONTHS - Consultation on the principle of "equal - equal" | q318 |  |
|  |  | Help from public organizations DURING 12 MONTHS - Advice on HIV / AIDS, sexually transmitted diseases | q316 |  |
| Any form of specialist medical or legal counselling | x | Help from public organizations DURING 12 MONTHS - Consultation of a psychologist | q319 | Receipt of specialist counselling = answered yes to q319 OR q320 |
|  |  | Help from public organizations DURING 12 MONTHS - Consultation of a lawyer | q320 |  |
| Online counselling | x | - | - | - |
| Some form of peer counselling | x | - | - | - |
| Any form of HIV testing and counselling | x | What kind of help or services did you receive from NGOs FOR THE LAST 12 MONTHS?_Free testing for HIV / AIDS | q313 | Receipt of HIV testing = answered yes to q313 |
| Participation in group work | x | Help from public organizations DURING 12 MONTHS - Support for mutual support / mutual assistance groups | q315 | Participation in group work = answered yes to q315 |
| Male condoms | x | What kind of help or services did you receive from NGOs FOR THE LAST 12 <MONTHS? _ Receiving condoms | q326 | Receipt of male condoms = answered yes to 1326 |
| Female condoms and training for usage |  |  |  |  |
| Lubricants | x | - | - | - |
| Some form of TB screening |  |  |  |  |
| Any form of STI testing | x | Have you passed the chlamydia test with the quick tests in 2012? | q408 | Receipt of any form of STI testing = yes to ANY of the following: q408, q413, q312, q406, q411, q407, q412, q296, q290, q260, q266, q284, q254, q311 |
|  |  | Have you passed the test for chlamydia using rapid tests in 2013? | q413 |  |
|  |  | Help from public organizations DURING 12 MONTHS - Free testing for venereal disease | q312 |  |
|  |  | Have you passed the syphilis test with the quick tests in 2012? | q406 |  |
|  |  | Have you passed the test for syphilis with the quick tests in 2013? | q411 |  |
|  |  | Have you passed the test for gonorrhoea with the quick tests in 2012? | q407 |  |
|  |  | Have you passed the test for gonorrhoea using rapid tests in 2013? | q412 |  |
|  |  | How was treated - Candidiasis - Appealed to a public organization | q296 |  |
|  |  | How did you treat this disease? TRICHOMONIASIS_I went to NGO | q290 |  |
|  |  | Did you treat - Genital herpes - Turned to a public organization | q260 |  |
|  |  | How was treated - Chlamydia - turned to a public organization | q266 |  |
|  |  | How was treated - Syphilis - turned to a public organization | q284 |  |
|  |  | How was treated - Gonorrhoea - turned to a public organization | q254 |  |
|  |  | What kind of help or services did you receive from NGOs FOR THE LAST 12 MONTHS?_Free treatment for sexually transmitted diseases | q311 |  |
| Any form of hepatitis C testing | x | Have you passed the Hepatitis C test with the quick tests in 2012? | q410 | Receipt of any form of hepatitis C testing = yes to at least one of the following: q410, q415, q314 |
|  |  | Did you pass the Hepatitis C test with the help of the quick tests in 2013? | q415 |  |
|  |  | Help from public organizations DURING 12 MONTHS - Free hepatitis C testing | q314 |  |
| Any form of hepatitis B testing | x | Have you passed the test for Hepatitis B by means of rapid tests in 2013? | q414 | Receipt of any hepatitis B testing = yes to either q414 or q409 |
|  |  | Have you passed the Hepatitis B test with the quick tests in 2012? | q409 |  |
| Referral to OST |  |  |  |  |
| ^+^Syringe exchange and/or delivery |  | Help from public organizations DURING 12 MONTHS - Exchange of syringes | q322 | Receipt of syringes = yes to q322 |
| Overdose prevention |  |  |  |  |
| Training about safe behaviour |  |  |  |  |
| Case Management | x | - | - | - |
| Counteracting violence | x | - | - | - |
| Distribution of general medication | x | - | - | - |
| Antiseptics | x | What kind of help or services did you receive from NGOs FOR THE LAST 12 MONTHS?_Recovery of disinfection solutions | q323 | Receipt of antiseptics = yes to q323 |
| Pregnancy tests | x | - | - | - |
| Any form of professional/ skilled work training | x | - | - | - |
| Organisation of leisure activities | x | - | - | - |
| Humanitarian aid |  |  |  |  |
| Basic household services |  |  |  |  |
| Tea and meals |  |  |  |  |
| Childcare and services | x | - | - | - |
| Cosmetologist and hairdresser services | x | - | - | - |
| ^+^Intimate hygiene items |  | Help from public organizations DURING 12 MONTHS - Obtaining the objects of hygiene | q325 | Receipt of hygiene items = yes to q325 |
| Referrals to relevant specialists | x | - | - | - |
| *Awareness raising and educational material | x | Help from public organizations DURING 12 MONTHS - Reception of information booklets, brochures | q324 | Receipt of awareness raising material = yes to q324 |
| **2015** | | | | |
| **PP component** | **Included in PP** | **Corresponding interview question in IBBS** | **Variable name** | **Definition of variable for analysis** |
| Any form of counselling from a social worker | x | - | - | - |
| Any form of specialist medical or legal counselling |  |  |  |  |
| Online counselling |  |  |  |  |
| Some form of peer counselling | x | - | - | - |
| Any form of HIV testing and counselling | x | Please tell me exactly where you applied for the HIV test? - To a non-governmental organization | q592 | Receipt of any form of HIV testing = yes to (q592 OR q648 OR q649) AND q622 |
|  |  | Let's clarify if it was during the LAST 12 MONTHS? | q622 |  |
|  |  | Did you pass HIV testing using quick tests in a public organization in 2014? | q648 |  |
|  |  | Did you pass HIV testing using quick tests in a public organization in 2015? | q649 |  |
| Participation in group work |  |  |  |  |
| Male condoms | x | Have you received the previous 12 months free condoms? | q501 | Receipt of male condoms = yes to q501 |
| Female condoms and training for usage |  |  |  |  |
| Lubricants | x | - | - | - |
| Some form of TB screening | x | - | - | - |
| Any form of STI testing | x | Have you been tested for such infections through rapid tests in a public organization during 2014: Chlamydia | q651 | STI testing = yes to q651 OR q656 OR q650 OR 655 OR q657 OR q 652 OR q451 OR q493 OR 481 OR q463 OR q457 OR q487 |
|  |  | Have you tested for such infections through rapid tests in a public organization during 2015: Chlamydia | q656 |  |
|  |  | Have you been tested for such infections through rapid tests in a public organization during 2014: Syphilis | q650 |  |
|  |  | Have you tested for such infections through rapid tests in a public organization during 2015: Syphilis | q655 |  |
|  |  | Have you tested for such infections through rapid tests in a public organization during 2015: Gonorrhoea | q657 |  |
|  |  | Have you been tested for such infections through rapid tests in a public organization during 2014: Gonorrhoea | q652 |  |
|  |  | How was treated - Gonorrhoea - turned to a public organization | q451 |  |
|  |  | How was treated - Candidiasis - Appealed to a public organization | q493 |  |
|  |  | How was treated - Syphilis - turned to a public organization | q481 |  |
|  |  | How was treated - Chlamydia - turned to a public organization | q463 |  |
|  |  | How did you treat this disease? - I turned to a public organization: Genital herpes | q457 |  |
|  |  | How did you treat this disease? TRICHOMONIASIS_I went to NGO | q487 |  |
| ^+^Any form of hepatitis C testing |  | Have you been tested for such infections through rapid tests in a public organization during 2014: Hepatitis C | q654 | Hepatitis C testing receipt = (Rapid test 2014) OR (rapid test 2015) |
|  |  | Have you been tested for such infections through rapid tests in a public organization during 2015: Hepatitis C | q659 |  |
| Any form of hepatitis B testing | x | Have you tested for such infections through rapid tests in a public organization during 2015: Hepatitis B | q658 | Hepatitis B testing receipt = (rapid test 2014=Y) OR (rapid test 2015=Y) |
|  |  | Have you tested for such infections through rapid tests in a public organization during 2014: Hepatitis B | q653 |  |
| Referral to OST | x | - | - | - |
| Syringe exchange and/or delivery |  |  |  |  |
| Overdose prevention |  |  |  |  |
| Training about safe behaviour |  |  |  |  |
| Case Management |  |  |  |  |
| ^+^Counteracting violence |  |  |  |  |
| Distribution of general medication |  |  |  |  |
| Antiseptics |  |  |  |  |
| Pregnancy tests |  |  |  |  |
| Any form of professional/ skilled work training |  |  |  |  |
| Organisation of leisure activities |  |  |  |  |
| Humanitarian aid |  |  |  |  |
| Basic household services |  |  |  |  |
| Tea and meals |  |  |  |  |
| Childcare and services |  |  |  |  |
| Cosmetologist and hairdresser services |  |  |  |  |
| Intimate hygiene items |  |  |  |  |
| Referrals to relevant specialists | x | - | - | - |
| *Awareness raising and educational material | x | - | - | - |
| **2017** | | | | |
| **PP component** | **Included in PP** | **Corresponding interview question in IBBS** | **Variable name** | **Definition of variable for analysis** |
| Any form of counselling from a social worker | x | - | - | - |
| Any form of specialist medical or legal counselling |  |  |  |  |
| Online counselling |  |  |  |  |
| Some form of peer counselling | x | - | - | - |
| Any form of HIV testing | x | Let's clarify if it was during the LAST 12 MONTHS? | q465 | Receipt of any form HIV testing = yes if: (q465 = yes) AND (q444 = yes) AND (q503 OR q504) |
|  |  | Did you make a fast-acting HIV test in a non-governmental organization (or mobile clinic, at home or on the street with the help of a social worker) in 2016? | q503 |  |
|  |  | Did you make a fast-acting HIV test in a non-governmental organization (or mobile clinic, at home or on the street with the help of a social worker) in 2017? | q504 |  |
|  |  | Please tell me exactly where you were tested for HIV? (Screening (primary) HIV test) - In a public organization / mobile clinic / on the street or at home through a social (outreach) worker | q444 |  |
| Participation in group work |  |  |  |  |
| Male condoms | x | Have you received condoms for free during the last 12 months (from a representative of a public organization, a health worker, at night clubs, at parties, etc.)? | q330 | Male condom receipt = Y = q330 |
| Female condoms and training for usage |  |  |  |  |
| Lubricants | x | - | - | - |
| Some form of TB screening | x | - | - | - |
| Any form of STI testing | x | - | - | - |
| ^+^Any form of hepatitis C testing |  | Did you make a hepatitis test C fast-paced test in a non-governmental organization in 2016? | q524 | Hepatitis C treatment receipt = (rapid test 2017 = Y) OR (rapid test 2018 = Y) |
|  |  | Did you make a hepatitis test C fast-paced test in a non-governmental organization in 2017? | q525 |  |
| Any form of hepatitis B testing | x | - | - | - |
| Referral to OST | x | - | - | - |
| Syringe exchange and/or delivery |  |  |  |  |
| Overdose prevention |  |  |  |  |
| Training about safe behaviour |  |  |  |  |
| ^+^Case Management |  |  |  |  |
| ^+^Counteracting violence |  |  |  |  |
| Distribution of general medication |  |  |  |  |
| Antiseptics |  |  |  |  |
| Pregnancy tests |  |  |  |  |
| Any form of professional/ skilled work training |  |  |  |  |
| Organisation of leisure activities |  |  |  |  |
| Humanitarian aid |  |  |  |  |
| Basic household services |  |  |  |  |
| Tea and meals |  |  |  |  |
| Childcare and services |  |  |  |  |
| Cosmetologist and hairdresser services |  |  |  |  |
| Intimate hygiene items |  |  |  |  |
| Referrals to relevant specialists | x | - | - | - |
| *Awareness raising and educational material | x | - | - | - |
| *OST = Opioid Substitution Therapy; TB = Tuberculosis* | | | | |
| ** Not explicitly listed in the PP components, however described in detailed in the 'prevention for FSW' section* | | | | |
| ^+^*Components not included in the general annual reports but included in the IBBS questionnaire.* | | | | |
