## Supplementary material for "Access to HIV-prevention in female sex workers in Ukraine between 2009 and 2017: coverage, barriers and facilitators": S5 Table. Independent variables from IBBS

### Supporting Information S5: Independent variables from IBBS and corresponding thematic clusters for 2009, 2011, 2013, 2015 and 2017

| **Thematic cluster** | **Specific risk factor** | **Question in IBBS** | **2009** | **2011** | **2013** | **2015** | **2017** | **Definition** |
| --- | --- | --- | --- | --- | --- | --- | --- | --- |
| Legal status of FSW | Criminalisation of FSW |  |  |  |  |  |  |  |
|  | Current incarceration |  |  |  |  |  |  |  |
|  | History of police arrest |  |  |  |  |  |  |  |
|  | Fear of arrest |  |  |  |  |  |  |  |
|  | Demolition of red-light districts |  |  |  |  |  |  |  |
|  | Infringement of HCF and NGOs who work with FSW |  |  |  |  |  |  |  |
| Phrasing of FSW law | Conflation of the term 'sex trafficking' with 'sex work' |  |  |  |  |  |  |  |
|  | Implementation of brothel rescue-raid police practices to combat sex trafficking |  |  |  |  |  |  |  |
| Legal support of key-populations | Existence of antidiscrimination laws |  |  |  |  |  |  |  |
|  | Violence against FSW formally recorded |  |  |  |  |  |  |  |
|  | Active monitoring and oversight of antidiscrimination policy |  |  |  |  |  |  |  |
| Political commitment towards reducing the HIV burden among key populations | Investment in HIV-related programmes for FSW (e.g. legal literacy) |  |  |  |  |  |  |  |
| Existence of institutions that exercise and enforce non-discriminatory practices (e.g. hiring of SW) | Presence of institutions that practice antidiscrimination policies (e.g. hiring of SW) |  |  |  |  |  |  |  |
| National prevalence of homelessness | Residential instability amongst FSW |  |  |  |  |  |  |  |
| FSW stigma amongst the general population | Fear of being identified as a FSW |  |  |  |  |  |  |  |
|  | High level of FSW stigma in the general population |  |  |  |  |  |  |  |
|  | Internationalised stigma from belief that all FSW are HIV positive | F9. Why have not you been tested for HIV for the LAST 12 MONTHS?_I do not want to know the results | p247_2 | V223_2 |  |  |  |  |
|  |  | If not, what do you see for yourself why not buy a HIV test in a pharmacy? - I do not want to do a HIV test at all |  |  | q393 | q584 | q514 |  |
| HIV stigma amongst the general population | Fear of stigma of being HIV positive | If not, what do you see for yourself why not buy a HIV test in a pharmacy? - I'm afraid someone will see me when I buy a test |  |  |  |  | q516 |  |
|  |  | Why are you personally not available for testing?_I am afraid of publicity about my status | p242_8 | V216_8 | q378 | q583 |  |  |
|  | Lack of visible advertising for HIV prevention centres |  |  |  |  |  |  |  |
|  | Lack of online advertising for HIV prevention centres |  |  |  |  |  |  |  |
|  | Fear of receiving a positive HIV test result |  |  |  |  |  |  |  |
|  | Exposure to anti-stigma interventions e.g. skits normalising FSW |  |  |  |  |  |  |  |
| Location of HIV prevention service centres | HCF is too far away from place of residence | F4. Why are you personally not available for testing?_Inconvenient location of the institution / point / centre of testing | P242_6 | V216_6 | q376 | q581 | q384 |  |
|  | Prevention facility is publicly visible |  |  |  |  |  |  |  |
|  | HIV prevention centre are *not* integrated in the HCF |  |  |  |  |  |  |  |
|  | Location of HCF: situated in local area |  |  |  |  |  |  |  |
| Regional mobility associated with FSW | High mobility of sex work |  |  |  |  |  |  |  |
|  | Short term travel to sex hot-spots | А5. Have you travelled from this city [CITY, WHERE YOU ARE RESEARCH] for more than 1 month [30 days] for the LAST 12 MONTHS for the purpose of providing sexual services? | p15 |  |  |  |  |  |
|  |  | Did you leave this city for more than 1 month [30 days] during the last 12 MONTHS in order to provide sexual services? |  | V17 | q37 | q58 | q39 |  |
|  | Intra-urban or intra-district mobility |  |  |  |  |  |  |  |
|  | Does not speak the local language |  |  |  |  |  |  |  |
|  | Ever having practiced SW in a foreign country |  |  |  |  |  |  |  |
|  | Not registered locally |  |  |  |  |  |  |  |
| Accepted behaviour of HCW | Lack of confidentiality exhibited by HCW |  |  |  |  |  |  |  |
|  | Distrust in HCW e.g. belief that HCW will administer the wrong drugs to deliberately kill a FSW |  |  |  |  |  |  |  |
|  | High level of stigdma exhibited by HCW | **Using a scale of 1 to 10**, please rate the following criteria that apply to your latest test with a quick test in a non-governmental organization? - Confidentiality |  |  |  |  | q511 | ≤ = dissatisfied |
|  |  | F4. Why are you personally not available for testing?_Dissatisfied with the attitude of the staff | p242_7 | V216_7 | q377 | q582 |  |  |
|  | High level of trust in HCW |  |  |  |  |  |  |  |
|  | History of discrimination at a HCF e.g. when disclosing sexual practices |  |  |  |  |  |  |  |
| HCF policies regarding the receipt of HIV prevention | Existence of policies that are insensitive to the FSW community e.g. Abstinence only, monogamous couples only, requirement to bring sexual partner, demand for identity card |  |  |  |  |  |  |  |
|  | Policies that neglect individual consent e.g. mandatory HIV testing policy |  |  |  |  |  |  |  |
| HIV prevention also available from NGOs | Poor quality of prevention services provided from NGOs e.g. distribution of condoms that break without the application of lubricant |  |  |  |  |  |  |  |
|  | Existence of NGOs that provide *voluntary* prevention services |  |  |  |  |  |  |  |
|  | Availability of telephone counselling at NGOs |  |  |  |  |  |  |  |
|  | Case management practiced at NGO |  |  |  |  |  |  |  |
|  | Existence of NGOs that provide *anonymous* prevention services |  |  |  |  |  |  |  |
| Functional hours of HCF | Appropriate opening hours of HCF e.g. 'moonlight' hours |  |  |  |  |  |  |  |
|  | Long waiting time at HCF | Using a scale of 1 to 10, please rate the following criteria that apply to your latest test with a quick test in a non-governmental organization? - A convenient time |  |  |  |  | q508 | inconvenient ≤5 |
|  |  | F4. Why are you personally not available for testing?_Inconvenient work schedule of institution / point / centre of testing | P242_5 | V216_5 | q375 | q580 |  |  |
| Available services at HCF | Availability of self-help services e.g. HIV self-tests |  |  |  |  |  |  |  |
|  | Perception that only low-quality services are available at HCF e.g. fear of being infected by medical testing utensils at the HCF |  |  |  |  |  |  |  |
|  | Lack of available services that are tailored to the FSW community e.g. no anal and vaginal examinations |  |  |  |  |  |  |  |
|  | A lack of harm reduction services offered at HCF e.g. OST |  |  |  |  |  |  |  |
|  | Insufficient supply of HIV prevention |  |  |  |  |  |  |  |
|  | Support services for HCW e.g. training about the specific needs of FSWs |  |  |  |  |  |  |  |
| National financial scheme for HIV prevention | Bribes are expected from HCW |  |  |  |  |  |  |  |
|  | Transportation to HCF is too expensive |  |  |  |  |  |  |  |
|  | HCF services are free of charge | F4. Why are you personally not available for testing? _ There is no money for testing | p242_4 | v216_4 | q374 | q579 |  |  |
|  |  | Why did this happen ( Tell me, please, have there been cases when you could not buy condoms when you needed them during the last 30 days?)? --> Condoms cost too much |  |  |  | q517 | q384 |  |
| Level of incentive for women to (re-) enter the workforce | FSW is ones sole source of income | А2. Which of the proposed types corresponds to your social status [if you do not take into account your employment in the field of commercial sex]? --> unemployed | p10 | V12 | q32 | q39 | q19 |  |
| National median household income | Higher annual household income (higher than average) |  |  |  |  |  |  |  |
|  | Currently attending a private doctor for HIV prevention |  |  |  |  |  |  |  |
| FSW involvement in the development of HIV prevention strategies | Prevention services are carefully tailored to the FSW community |  |  |  |  |  |  |  |
|  | History of participation in a HIV program/intervention |  |  |  |  |  |  |  |
| FSW engagement with government | Existing dialogue between FSW community and government |  |  |  |  |  |  |  |
| FSW engagement with police | Existing dialogue between FSW community and police e.g. training conducted by/with FSW for police about supporting (at least not impeding) FSWs access to health care |  |  |  |  |  |  |  |
| Existence of FSW community organisations (e.g. NGOs or CBOs run by or involving FSWs) | Membership in an NGO that works with FSW community | Are you a client of an organization that deals with HIV prevention among sex workers, that is, do you have a plastic card customer, through which you receive condoms or other services from social workers? |  |  |  |  | q331 |  |
|  |  | D3.1 Are you a client of any public organization (have a card or an individual code) that works with commercial sex women or injecting drug users? | p206 | V190 | q305 | q502 |  |  |
| Collaboration and cooperation between groups of FSW and non-FSW | Participation in non-sex worker community organisation or social networks |  |  |  |  |  |  |  |
| Existence of peer-education programs | Exposure to peer- education |  |  |  |  |  |  |  |
| Existence of prevention services at work place | Availability of condoms and lubricant at work place |  |  |  |  |  |  |  |
| Soliciting setting | Village-based soliciting |  |  |  |  |  |  |  |
|  | Street-based soliciting | How did you usually find / met / searched your clients for the LAST MONTH (30 days)? - On the street (open area, park, squares, etc.) | p25 | V27 | q115 | q293 | q196 |  |
|  | Working in venues with less client traffic |  |  |  |  |  |  |  |
|  | Highway-based soliciting ^1^ | Please tell me among the following ways to find clients that you consider to be BASIC **(standard)** to yourself? --> On the highway | p25 | V27 | q115 | q293 | q196 |  |
|  | Lodge/hotel-based soliciting | Please tell me among the following ways to find clients that you consider to be BASIC (standard) to yourself? --> The hotel / motel | p25 | V27 | q115 | q293 | q196 |  |
|  | Home-based soliciting |  |  |  |  |  |  |  |
|  | Affluent entertainment venues soliciting | Please tell me among the following ways to find clients that you consider to be BASIC **(standard)** to yourself? -->At the casino, club, bar, disco, etc. | p25 | V27 | q115 | q293 | q196 |  |
| Local police practices | Pervasive patrolling of HIV facilities |  |  |  |  |  |  |  |
| Level of gender-based violence at work place | Experience of gender-based violence from anyone | Have you been subjected to violence (such as beatings, rape, verbal humiliation, extortion, etc.) during the provision of sex services? |  | V211 | q339 | q541 | q404 |  |
|  | Experience of gender-based violence from police officer in order to avoid arrest | Who hurt violence? - An employee of law enforcement agencies |  | V212_3 | q353 | q557 | q420 |  |
|  | Experience of gender-based violence from client | Who hurt violence? - Customers |  | V212_1 | q350 | q554 | q417 |  |
| Venue client policies | Client sign in policy |  |  |  |  |  |  |  |
|  | Removal of violent clients |  |  |  |  |  |  |  |
| Family history of HIV | Being an orphaned child from a parent who has died from HIV |  |  |  |  |  |  |  |
|  | Experience of family living with or dying from HIV |  |  |  |  |  |  |  |
| Number of clients | High number of clients | В5. How many different CLIENTS, whom you provided sexual services for remuneration, did you have FOR THE LAST WORKING DAY (24 HOURS)? | p36 | V40 | q123 | q308 | q206 |  |
| Condom use | Consistent condom use with casual clients within the past 6 months |  |  |  |  |  |  |  |
|  | Pressure for sex without a condom from client | В8. Why did not you use a condom during sexual intercourse with your LAST client?_The client insisted on not using a condom | p39_5 | V44_5 | q130 | q320 | q216 |  |
| Gender-violence | Experience of gender-based violence from an intimate partner | Who hurt violence? - Permanent sexual partner |  | v212_2 | q351 | q555 | q418 |  |
| Condom use | Consistent condom use with a boyfriend or lover within the past 6 months |  |  |  |  |  |  |  |
| Partnership status | Living with partner | From the options, choose the one that matches your family status at the moment --Married and live with my husband | p18 | V20 | q38 | q190 | q138 | Yes to one of these |
|  |  | From the options, choose the one that matches your family status at the moment -- Married, but I live with another sexual partner | p18 | V20 | q38 | q190 | q138 |  |
|  |  | From the options, choose the one that matches your family status at the moment --Officially unmarried, but live with a regular partner | p18 | V20 | q38 | q190 | q138 |  |
|  | Not living with a partner | From the options, choose the one that matches your family status at the moment -- Married, but do not live together with anyone | p18 | V20 | q38 | q190 | q138 | Yes to either of these |
|  |  | From the options, choose the one that matches your family status at the moment -- Unmarried, I do not live with a sexual partner | p18 | V20 | q38 | q190 | q138 |  |
| Level of awareness about FSW status | Non-paying partner is aware of FSW status | G2.1 How many of these people know that you are providing sex services for a fee? MEMBERS OF YOUR FAMILY | p255 |  |  |  |  |  |
|  |  | Does your husband or partner with whom you live know that you are providing sex services for rewards? |  | q243 | q39 | q191 | q139 |  |
| Level of education | Junior high school or lower level of education | Your education | p9 | V11 | q31 | q38 | q18 |  |
| Work life balance | Lack of time to seek health care | How many days did you provide sexual services for a fee during the last week [7 days]? |  |  |  | q202 | q146 |  |
|  |  | How many days do you work for this week? | p22 | V24 | q68 |  |  |  |
| History of substance abuse | History of binge drinking |  |  |  |  |  |  |  |
|  | Any history of illicit drug use |  |  |  |  |  |  |  |
|  | Indifferent outlook to life due to drug addiction, thus not caring about health and wellbeing |  |  |  |  |  |  |  |
|  | History of sharing needles | Did you use a sterile needle and syringe during the last injection? |  |  |  | q437 | q301 |  |
|  |  | С4. Did you use a joint injection tool (syringe, a needle that someone else used to use) during the LAST INJECTION? | p129 | V143 | q236 |  |  |  |
| Duration in sex work | FSW begin | How old were you when you first provided sexual services for a reward (got money or otherwise)? | p27 | V31 | q71 | q205 | q149 |  |
|  | Sexual debut | Age of sexual debut | p26 | V30 | q70 | q204 | q148 |  |
|  | Duration working as a FSW longer than 1 year |  |  |  |  |  |  |  |
| Knowledge of HIV | Knowledge of where to get a free HIV test | Do you know where to go if you want to take an HIV test? | p239 | V214 | q369 | q574 | q438 |  |
|  | Knowledge of HIV transmission routes | How much do you agree with the following statements about HIV: HIV infection can be avoided if you have sex with only one loyal non-infected partner | p223 | V200 | q329 | q531 | q396 | Knowledge = all five correct |
|  |  | How do you agree with the following statements about HIV: HIV infection can be avoided if you correctly use a condom during each sexual intercourse | p224 | V201 | q330 | q532 | q397 |  |
|  |  | How do you agree with the following statements about HIV: A person can become infected with HIV if he / she drank one glass of HIV-infected person | p227 | V204 | q333 | q535 | q399 |  |
|  |  | How do you agree with the following statements about HIV: A person can become infected with HIV through sharing with an HIV-infected person a toilet, a swimming pool, a sauna | p228 | V205 | q334 | q536 | q400 |  |
|  |  | How much do you agree with the following statements about HIV: HIV infection can be infected using a needle for injection that was used by another person | p229 | V206 | q335 | q537 | q401 |  |
|  | Belief that prophylactic use of antibiotics can prevent HIV acquisition |  |  |  |  |  |  |  |
|  | Belief that FSW can detect clients with symptoms of infection |  |  |  |  |  |  |  |
|  | Self perception of being high risk for HIV |  |  |  |  |  |  |  |
| HIV factors | HIV positive status | F13.1 HIV status --> yes | p252 | V228 | q401 | q627 | q471 |  |
|  | History of side effects from HIV medication |  |  |  |  |  |  |  |
|  | History of being diagnosed with HIV during the first visit to a prevention centre |  |  |  |  |  |  |  |
| General health status | Unresolved clinical symptoms |  |  |  |  |  |  |  |
| STI factors | Hepatitis C status ^2^ | D1.1.6 Have you had these diseases for the LAST 12 MONTHS? HEPATITIS С | p170 | V174 | q274 | q471 | q327 |  |
|  | Hepatitis B status ^2^ | D1.1.5 Have you had these diseases for the LAST 12 MONTHS? HEPATITIS B | p163 | V171 | q268 | q465 | q324 |  |
|  | Positive gonorrhoea status |  |  |  |  |  |  |  |
| Reproductive status | Motherhood | А9. Are there people [children, parents, friends, acquaintances, etc.], which you hold at the expense of your earnings | p19 | V21 | q54 | q193 | q141 |  |
|  | Number of dependents^1^ | Number - The child / children |  |  | q56 | q195 | q142 | Sum of total |
|  |  | Number - husband / cohabitant |  |  | q58 | q197 | q143 |  |
|  |  | Number - parents / grandma / grandpa |  |  | q60 | q199 | q144 |  |
|  |  | Number - friends |  |  | q62 |  |  |  |
|  |  | Number - familiar |  |  | q64 |  |  |  |
|  |  | Number - Someone else |  |  | q66 | q201 | q145 |  |
|  |  | Number - general |  |  | q67 |  |  |  |
|  |  | А10. How many people, without you considering you, hold on account of your earnings? | p20 | V22 |  |  |  |  |
|  | Pregnancy |  |  |  |  |  |  |  |
| Age | Age between 18-24 | That is to you now (age?) | p8 | V10 | q15 | q33 | q15 |  |
| Age client | Younger Clientele | And among the age groups you mentioned, which representatives most likely to meet you most for the LAST month (30 days)? (Age group) --> Teens under 18 | p31 | V35 | q89 | q266 | q169 |  |
|  |  | And among the age groups you mentioned, which representatives most likely to meet you most for the LAST month (30 days)? (Age group) --> Youth 18-25 | p31 | V33 | q89 | q266 | q169 |  |
|  | Older clientele | And among the age groups you mentioned, which representatives most likely to meet you most for the LAST month (30 days)? (Age group) -->Men of middle age (36-50 years) | p31 | V33 | q89 | q266 | q169 |  |
|  |  | And among the age groups you mentioned, which representatives most likely to meet you most for the LAST month (30 days)? (Age group) --> Men are over 50 yrs. old | p31 | V33 | q89 | q266 | q169 |  |
| Subset question | Belief that HIV is personally not available | F3. Is HIV testing available to you? | p241 | V215 | q370 | 575 | q513 |  |
|  | Condom use | Did you use a condom last time? | p37 | V41 | 124 | 313 | 209 |  |
| ^1^Variable not identified during the literature review, but available in the IBBS  ^2^ Variable included in the IBBS but not explicitly listed in the PP | | | | | | | | |
