## Supplementary material for "Access to HIV-prevention in female sex workers in Ukraine between 2009 and 2017: coverage, barriers and facilitators": S6 Table. Receipt or non-receipt of extended PP

### Supporting Information S6: Barriers and facilitators to accessing HIV prevention stratified by receipt or non-receipt of extended PP between 2009 and 2017

|  |  | 2009 (%) | 2011 (%) | 2013 (%) | 2015 (%) | 2017 (%) |
| --- | --- | --- | --- | --- | --- | --- |
| Macrostructural | | | | | | |
| Socio-cultural | | | | | | |
| *Level of HIV stigma amongst the population* | | | | | | |
| Belief that HIV testing is personally *not* accessible | Received | 1.31 | 1.52 | 0.43 | 0.58 | 24.83 |
|  | Not Received | 12.54 | 10.35 | 7.15 | 4.13 | 25.23 |
| Fear of HIV status being exposed^1^ | Received^1^ | 40.00^1^ | 20.00^1^ | 0.12^1^ | 0.04^1^ | 1.30 |
|  | Not Received^1^ | 31.05^1^ | 16.76^1^ | 1.02^1^ | 0.69^1^ | 3.91 |
| No desire to know HIV test result^1^ | Received^1^ | 9.89^1^ | 10.36^1^ | 0.86^1^ | - | 0.49 |
|  | Not Received^1^ | 6.67^1^ | 10.81^1^ | 0.83^1^ | 0.21^1^ | 2.89 |
| Geographical | | | | | | |
| *Location of HIV prevention service centres* | | | | | | |
| Inconvenient location of prevention service centres^1^ | Received^1^ | 6.67^1^ | 4.21^1^ | 0.06^1^ | - | 0.80 |
|  | Not Received^1^ | 0.81^1^ | 4.4^1^ | 0.7^1^ | 0.16^1^ | 1.29 |
| *Migration associated with FSW* | | | | | | |
| History of travel for FSW purposes | Received | 20.41 | 15.18 | 12.56 | 8.17 | 7.72 |
|  | Not Received | 14.54 | 14.99 | 17.62 | 7.04 | 10.66 |
| Health-Related Policy | | | | | | |
| *Tolerated behaviour of health care staff* | | | | | | |
| Dissatisfaction with staff attitude^1^ | Received^1^ | 8.89^1^ | 8.42^1^ | 0.03^1^ | 0.04^1^ | 0.31 |
|  | Not Received^1^ | 2.82^1^ | 7.69^1^ | 1.60^1^ | 0.16^1^ | 0.12 |
| *Functional hours of health care facility* | | | | | | |
| Inappropriate opening hours^1^ | Received^1^ | 8.89^1^ | 7.37^1^ | 0.03^1^ | 0.04^1^ | 1.36 |
|  | Not Received^1^ | 1.21^1^ | 6.32^1^ | 0.38^1^ | 0.05^1^ | 0.61 |
| Economic | | | | | | |
| *National financial scheme for HIV prevention* | | | | | | |
| Belief that HIV prevention too costly^1^ | Received^1^ | 4.44^1^ | 13.68^1^ | 0.09^1^ | 2.33^1^ | 1.17 |
|  | Not Received^1^ | 20.56^1^ | 16.21^1^ | 0.64^1^ | 2.33^1^ | 2.60 |
| *National support schemas for women in the workforce* | | | | | | |
| FSW sole employment | Received | 38.05 | 46.52 | 41.94 | 58.29 | 60.41 |
|  | Not Received | 33.56 | 42.37 | 38.7 | 39.83 | 50.93 |
| Community Organisation - extended | | | | | | |
| Sex Worker Collectivisation | | | | | | |
| *Collective agency of FSW community* | | | | | | |
| Client of an NGO | Received | 67.90 | 82.12 | 89.69 | 92.16 | 90.24 |
|  | Not Received | 2.22 | 4.06 | 2.23 | 17.69 | 10.89 |
| Work Environment | | | | | | |
| Physical | | | | | | |
| *Soliciting setting* | | | | | | |
| Street | Received | 19.90 | 26.14 | 14.84 | 15.6 | 16.68 |
|  | Not Received | 9.77 | 14.57 | 15.3 | 9.72 | 8.50 |
| Highway | Received | 31.08 | 22.83 | 27.61 | 25.08 | 18.96 |
|  | Not Received | 19.76 | 14.89 | 16.2 | 15.79 | 7.80 |
| Hotel | Received | 6.46 | 3.94 | 3.81 | 3.25 | 5.31 |
|  | Not Received | 5.44 | 3.68 | 2.82 | 2.09 | 6.07 |
| Affluent settings | Received | 12.78 | 14.86 | 3.01 | 12.69 | 11.18 |
|  | Not Received | 25.86 | 17.13 | 11.52 | 17.35 | 20.53 |
| Social | | | | | | |
| *Gender violence at workplace* | | | | | | |
| History of gender violence whilst providing sexual services | Received | - | 50.46 | 51.17 | 50.66 | 44.41 |
|  | Not Received | - | 38.47 | 46.17 | 37.02 | 37.38 |
| Experience of gender violence from client^2^ | Received^2^ | - | 74.76^2^ | 45.09^2^ | 42.25^2^ | 35.52^2^ |
|  | Not Received^2^ | - | 71.71^2^ | 41.51^2^ | 29.98^2^ | 28.91^2^ |
| Experience of gender violence from a police officer^2^ | Received^2^ | - | 30.90^2^ | 11.64^2^ | 6.92^2^ | 8.09^2^ |
|  | Not Received^2^ | - | 23.02^2^ | 9.13^2^ | 5.51^2^ | 7.51^2^ |
| Interpersonal dynamic | | | | | | |
| Sex worker- client | | | | | | |
| *Condom use* | | | | | | |
| Condom usage during last sexual act | Received | 4.29 | 6.54 | 1.76 | 4.31 | 3.27 |
|  | Not Received | 13.82 | 15.32 | 5.87 | 9.38 | 6.92 |
| Pressure from client for sex without a condom | Received | 25.56 | 50.00 | 0.74 | 1.66 | 1.54 |
|  | Not Received | 36.23 | 34.07 | 2.43 | 4.56 | 3.48 |
| *Number of clients* |  |  |  |  |  |  |
| Number of clients in last 24 hours^+^ | Received^+^ | 2.06^+^ | 2.13^+^ | 2.10^+^ | 1.84^+^ | 2.44^+^ |
|  | Not Received^+^ | 1.60^+^ | 1.49^+^ | 1.79^+^ | 1.59^+^ | 2.18^+^ |
| Sex worker- partner | | | | | | |
| *Partner's knowledge of occupational status* | | | | | | |
| Partner knowledge of FSW status | Received | 38.79 | 36.65 | 17.72 | 18.62 | 15.87 |
|  | Not Received | 34.94 | 30.90 | 10.22 | 10.17 | 7.71 |
| *Gender-based violence* | | | | | | |
| Experience of gender violence from intimate partner | Received | - | 10.57 | 3.98 | 2.40 | 3.21 |
|  | Not Received | - | 15.40 | 3.45 | 1.64 | 3.04 |
| *Partnership status* | | | | | | |
| Living with a partner | Received | 34.28 | 27.50 | 28.86 | 33.83 | 31.07 |
|  | Not Received | 30.74 | 22.63 | 20.88 | 27.33 | 22.75 |
| Sex Worker Individual | | | | | | |
| Behavioural | | | | | | |
| *Knowledge of HIV* | | | | | | |
| Knowledge of HIV transmission routes (all questions correct) | Received | 63.69 | 63.45 | 58.92 | 57.75 | 58.18 |
|  | Not Received | 51.83 | 50.43 | 55.94 | 59.27 | 49.68 |
| Knowledge of where to access HIV prevention | Received | 96.80 | 97.43 | 99.1 | 98.96 | 98.58 |
|  | Not Received | 75.69 | 81.06 | 86.85 | 89.19 | 85.51 |
| *History of substance abuse* | | | | | | |
| Drug Injecting |  | - | - | - | - | - |
|  |  | - | - | - | - | - |
| History of sharing needles | Received | 22.55 | 8.19 | 1.88 | 13.31 | 9.94 |
|  | Not Received | 10.42 | 22.95 | 1.09 | 4.56 | 3.77 |
| *Work life balance* | | | | | | |
| Workdays per week^+^ | Received^+^ | 3.75^+^ | 3.98^+^ | 4.34^+^ | 4.43^+^ | 4.22^+^ |
|  | Not Received^+^ | 2.94^+^ | 3.05^+^ | 3.90^+^ | 3.66^+^ | 3.86^+^ |
| *Duration in sex work* | | | | | | |
| *Age of FSW debut*^+^ | Received^+^ | 20.52^+^ | 21.10^+^ | 21.74^+^ | 21.81^+^ | 23.01^+^ |
|  | Not Received^+^ | 20.38^+^ | 21.25^+^ | 21.10^+^ | 21.07^+^ | 22.05^+^ |
| *Age of sexual debut* | Received^+^ | 15.93^+^ | 15.84^+^ | 15.85^+^ | - | 16.40^+^ |
|  | Not Received^+^ | 15.98^+^ | 16.11^+^ | 15.82^+^ | - | 16.15^+^ |
| *Level of education* | | | | | | |
| Higher education | Received | 21.16 | 21.44 | 24.46 | 24.21 | 22.48 |
|  | Not Received | 19.89 | 24.92 | 17.5 | 28.15 | 30.01 |
| Biological | | | | | | |
| *HIV factors* | | | | | | |
| HIV positive status | Received | 12.88 | 8.91 | 3.7 | 3.86 | 3.27 |
|  | Not Received | 6.76 | 6.72 | 0.64 | 2.22 | 1.90 |
| *STI factors* | | | | | | |
| Hepatitis C within the last 12 months | Received | 6.03 | 6.38 | 8.61 | 7.59 | 8.28 |
|  | Not Received | 3.55 | 2.77 | 3.19 | 3.5 | 2.72 |
| Hepatitis B within the last 12 months | Received | 2.61 | 1.91 | 2.25 | 2.45 | 2.72 |
|  | Not Received | 1.33 | 0.69 | 1.21 | 1.11 | 2.22 |
| *Age* | | | | | |  |
| Age | Received^+^ | 27.90^+^ | 27.90^+^ | 29.03^+^ | 29.84^+^ | 31.61^+^ |
|  | Not Received^+^ | 26.59^+^ | 27.22^+^ | 27.42^+^ | 27.64^+^ | 28.69^+^ |
| *Reproductive status* | | | | | | |
| Presence of dependents | Received | 49.46 | 58.4 | 43.3 | 43.66 | 47.56 |
|  | Not Received | 48.06 | 49.31 | 56.32 | 55.14 | 58.38 |
| Number of dependents | Received^+^ | 1.92^+^ | 1.84^+^ | 1.88^+^ | 1.04^+^ | 0.93^+^ |
|  | Not Received^+^ | 1.79^+^ | 1.78^+^ | 1.73^+^ | 0.76^+^ | 0.58^+^ |
| Client individual | | | | | | |
| Biological | | | | | | |
| Age (≤ 35 years) | Received | 37.62 | 37.35 | 43.9 | 34.92 | 29.59 |
|  | Not Received | 44.28 | 41.30 | 43.64 | 43.89 | 34.20 |
| ^1^ Sub-question of “I believe that HIV testing is not available for me personally”  ^2^ Sub-question of “History of gender violence whilst providing sexual services’  ^+^ Mean | | | | | | |
